# Machine learning based predictive model of early mortality in stage III and IV prostate cancer

**DOI:** 10.1101/2020.04.15.20065938

**Authors:** Robert Chen

## Abstract

Prostate cancer remains the third highest cause of cancer-related deaths. Metastatic prostate cancer could yield poor prognosis, however there is limited work on predictive models for clinical decision support in stage III and IV prostate cancer.

We developed a machine learning model for predicting early mortality in prostate cancer (survival less than 21 months after initial diagnosis). A cohort of 10,303 patients was extracted from the Surveillance, Epidemiology and End Results (SEER) program. Features were constructed in several domains including demographics, histology of primary tumor, and metastatic sites. Feature selection was performed followed by regularized logistic regression. The model was evaluated using 5-fold cross validation and achieved 75.2% accuracy with AUC 0.649. Of the 19 most predictive features, all of them were validated to be clinically meaningful for prediction of early mortality.

Our study serves as a framework for prediction of early mortality in patients with stage II and stage IV prostate cancer, and can be generalized to predictive modeling problems for other relevant clinical endpoints. Future work should involve integration of other data sources such as electronic health record and genomic or metabolomic data.

## Introduction

Prostate cancer is the cancer with highest prevalence among men and remains the third highest cause of cancer-related deaths.[1] The median 5-year survival across all patients with prostate cancer is over 98%. However, only 30% of patients with advanced cancer have 5-year survival rate. The median overall survival of patients with distant prostate cancer is only 31%. [2]

There is limited work on the capabilities of predictive modeling approaches for early detection of mortality risk in patients with distant prostate cancer. One approach sought to stratify patients into subgroups with Cox multivariable regression [3], but used a limited feature set including solely clinical stage and Gleason score. Furthermore, while the prostate specific antigen [4]biomarker has been used in practice clinically as a prognostic marker, there is limited evidence of its predictive value due to low specificity.

Meanwhile, machine learning has shown to have strong potential for early detection of clinical endpoints in applications such as disease prediction[5–8], readmission prediction [9–11], drug adverse event prediction [12], among others.

In this study we developed a machine learning model based on logistic regression for prediction of early mortality from retrospective real-world data and evaluated their performance. Our model leverages predictive features in a variety of domains including demographics, histology, staging, tumor spread and metastatic status.

## Methods

A cohort of patients was selected from the The Surveillance, Epidemiology and End Results (SEER) program public retrospective dataset[13].

### Cohort Construction

Patients were selected from The Surveillance, Epidemiology and End Results (SEER) program was used to identify a cohort of 51,354 male patients who were diagnosed with prostate cancer between 2010 and 2015. Of these patients, inclusion and exclusion criteria were applied, resulting in a cohort of 10,303 patients to be used in the machine learning model.

#### Inclusion criteria

Patients were included if they had the following associated ICD-9 codes for prostate cancer: C619. The patient is required to have a group stage of III or IV. Furthermore, patients are included if they have a date of diagnosis between 2010 and 2015.

#### Exclusion criteria

Patients are excluded if they did not have follow-up information following their initial diagnosis.

Table 1 shows descriptive statistics of the cohort.

**Table 1:**
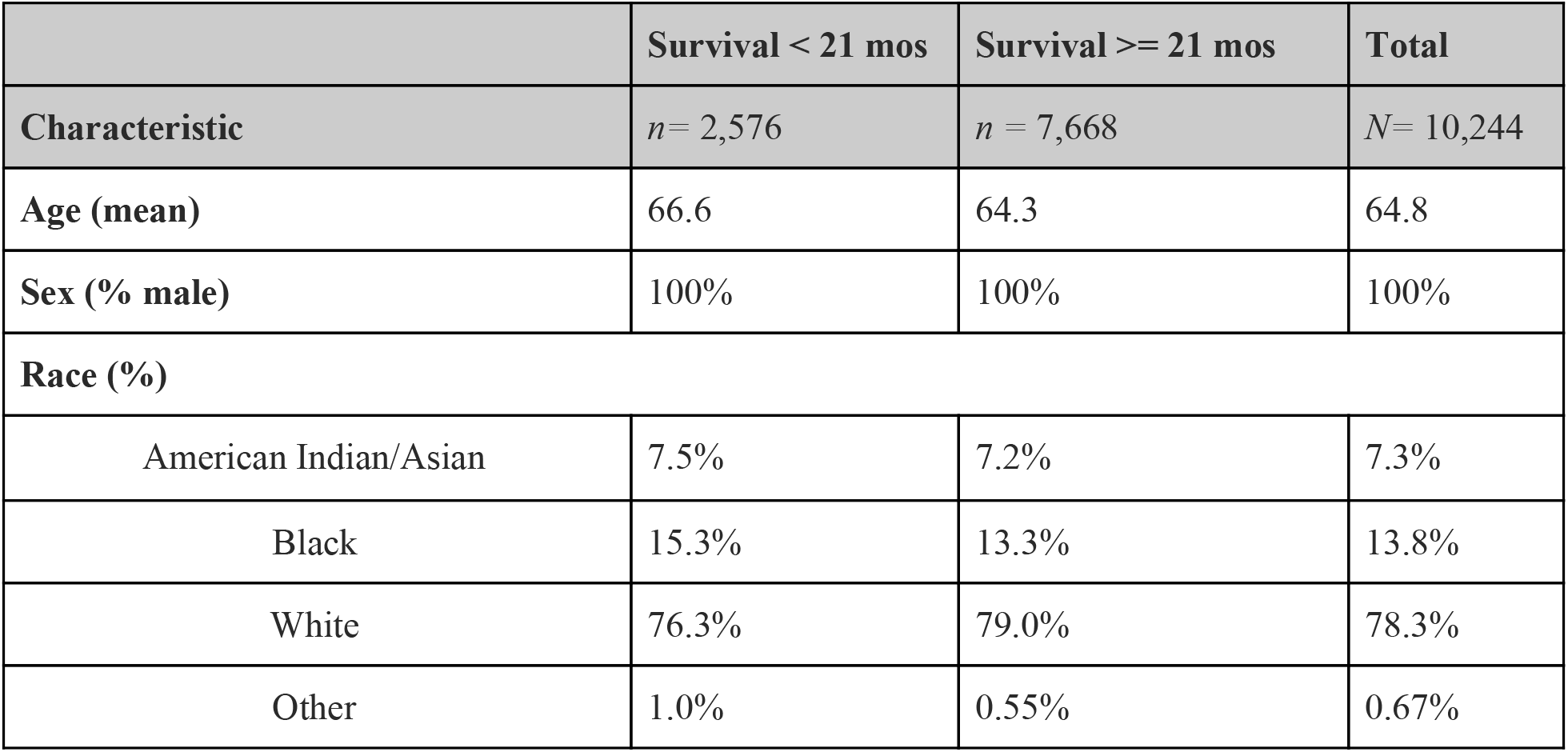
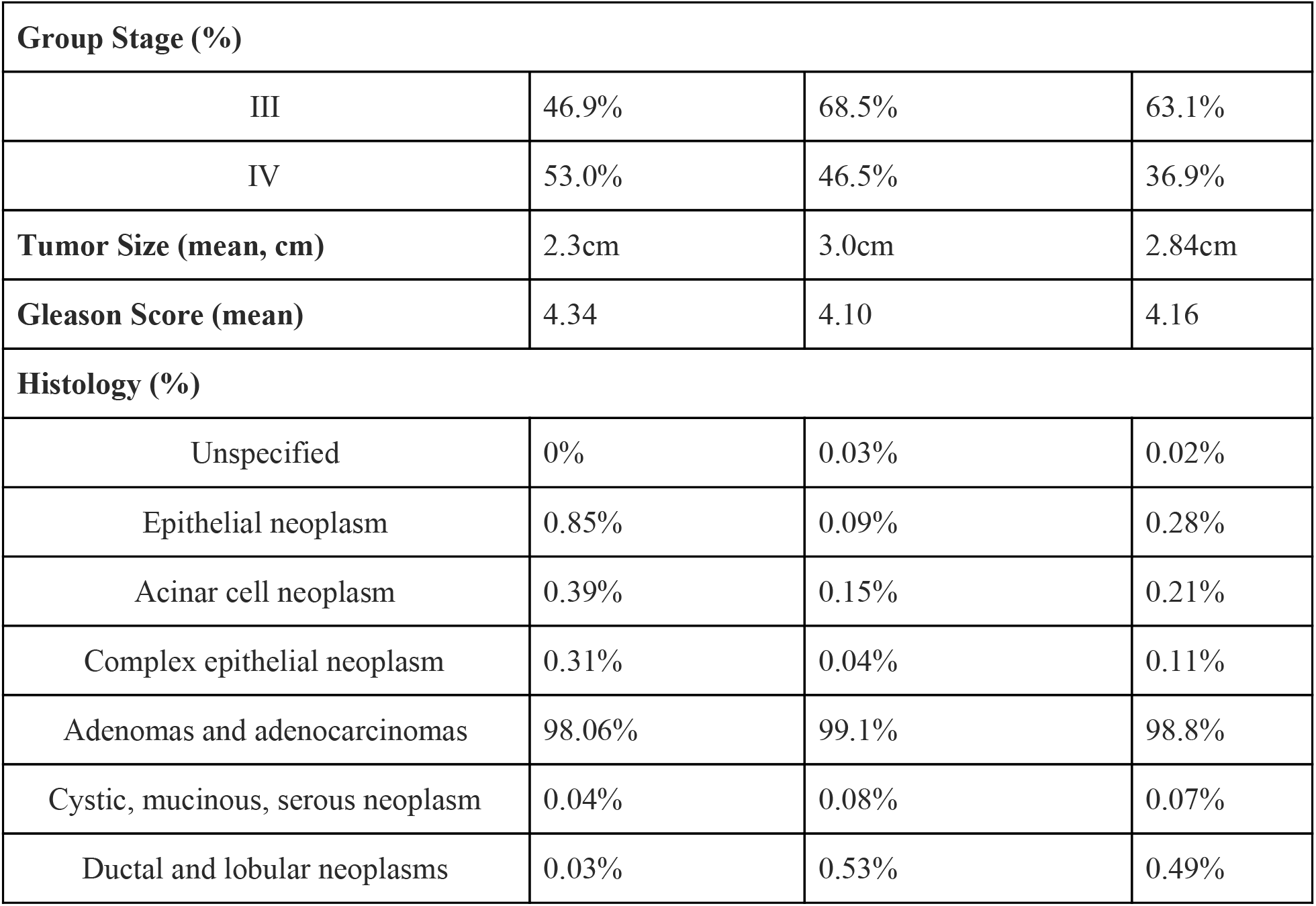
Baseline characteristics of the study cohort.

|  | Survival < 21 mos | Survival >= 21 mos | Total |
| --- | --- | --- | --- |
| Characteristic | *n*= 2,576 | *n* = 7,668 | *N*= 10,244 |
| Age (mean) | 66.6 | 64.3 | 64.8 |
| Sex (% male) | 100% | 100% | 100% |
| **Race (%)** |  |  |  |
| American Indian/Asian | 7.5% | 7.2% | 7.3% |
| Black | 15.3% | 13.3% | 13.8% |
| White | 76.3% | 79.0% | 78.3% |
| Other | 1.0% | 0.55% | 0.67% |

**Table 1:** Baseline characteristics of the study cohort.
| Group Stage (%) |  |  |  |
| --- | --- | --- | --- |
| III | 46.9% | 68.5% | 63.1% |
| IV | 53.0% | 46.5% | 36.9% |
| **Tumor Size (mean, cm)** | 2.3cm | 3.0cm | 2.84cm |
| **Gleason Score (mean)** | 4.34 | 4.10 | 4.16 |
| Histology (%) |  |  |  |
| Unspecified | 0% | 0.03% | 0.02% |
| Epithelial neoplasm | 0.85% | 0.09% | 0.28% |
| Acinar cell neoplasm | 0.39% | 0.15% | 0.21% |
| Complex epithelial neoplasm | 0.31% | 0.04% | 0.11% |
| Adenomas and adenocarcinomas | 98.06% | 99.1% | 98.8% |
| Cystic, mucinous, serous neoplasm | 0.04% | 0.08% | 0.07% |
| Ductal and lobular neoplasms | 0.03% | 0.53% | 0.49% |

Features corresponding to several domains were extracted for the cohort. These include demographics, AJCC staging criteria, metastatic sites, and prostate-specific criteria including Gleason score and biopsy-related findings. Table 2 shows the description of features used in the model.

**Table 2:** Features used in the model, as well as aggregation used in data preprocessing before model training.

| Feature | No. of features | Example | Aggregation |
| --- | --- | --- | --- |
| Age | 1 | 76.4 | Continuous |
| Race (White, Black, American Indian/Asian, Other) | 3 | Black | Categorical |
| Race - Hispanic | 1 | Hispanic | Categorical |
| Group Stage | 2 | III | Categorical |
| AJCC staging (T,N,M) | 19 | N0 | Categorical |
| Gleason Score | 2 | Score on Needle Core Biopsy | Continuous |
| Histology | 7 | Epithelial neoplasm | Categorical |
| Percentage of positive cores on biopsy | 1 | 30% | Continuous |
| Metastatic sites | 4 | Metastasis to Bone | Categorical |
| Tumor Size | 1 | 1.5 cm | Continuous |
| Lymph Node Involvement<sup>1</sup> | 8 | Regional lymph nodes removed for examination with pre-surgical systemic treatment or radiation, but lymph node evaluation based on clinical evidence. | Categorical |
| AJCC Metastasis Eval<sup>2</sup> | 7 | Meets criteria for AJCC pathologic staging of distant metastasi | Categorical |

### Kaplan Meier Analysis

A Kaplan Meier[14] analysis was performed with all patients in the cohort. The 25th percentile of overall survival time in months, was used as a threshold to determine class labels of patients:

0: patient survived at least the 25th percentile of overall survival
1: patient death occurred before the 25th percentile of overall survival

### Machine Learning Model

#### Feature Construction

Categorical features were one-hot encoded into separate features. For example, the feature *race* which includes categories (*white, black, other*) would be one hot encoded for a patient as (1,0,0) for *white*, (0,1,0) for *black*, and (0,0,1) for *other*. Continuous features were used in the current form. Standardization was performed on all features.

#### Predictive Modeling

Principal component analysis was performed on the cohort to reduce dimensionality. Feature selection was performed using ANOVA F-value for the. A logistic regression[15] model was trained using the features constructed, with the target labels

0: patient survived at least the 25th percentile of overall survival
1: patient death occurred before the 25th percentile of overall survival.

Grid search was performed to learn the most optimal set of modeling parameters from the following set: number of features {all}, regularization {*l1, l2*}, C {1e-2, 1e-1, 1, 1e1, 1e2}. The model was evaluated via 5-fold cross validation. The scikit-learn [16] Python package was used to implement the analysis.

## Results

### Kaplan Meier Analysis

The median overall survival was 37 months. The 25th percentile of survival time was 21 months, which was used in the definition of patient classes for the machine learning problem (Figure 1).

**Figure 1:**
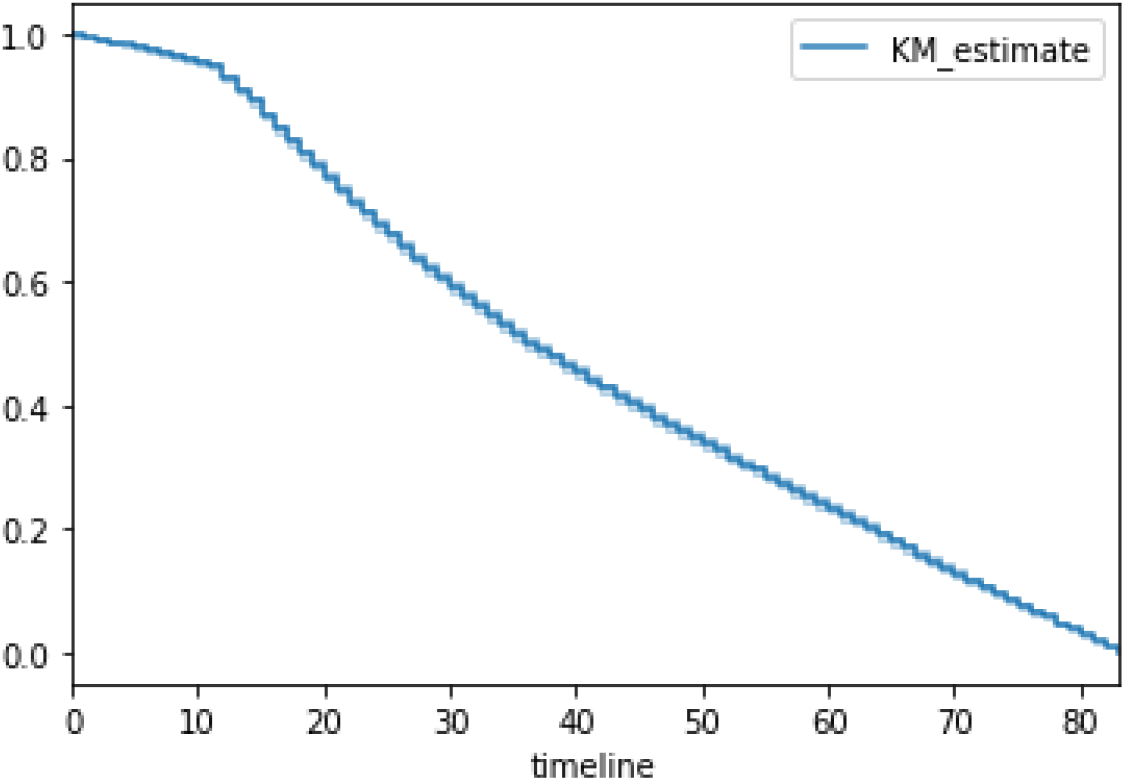
Kaplan Meier survival curve of all patients in the analytical cohort.

### Principal Component Analysis

We utilized principal components analysis (PCA) to visualize the variation in the cohort of patients. Figure 2 shows the patients projected onto the first two principal components.

**Figure 2:**
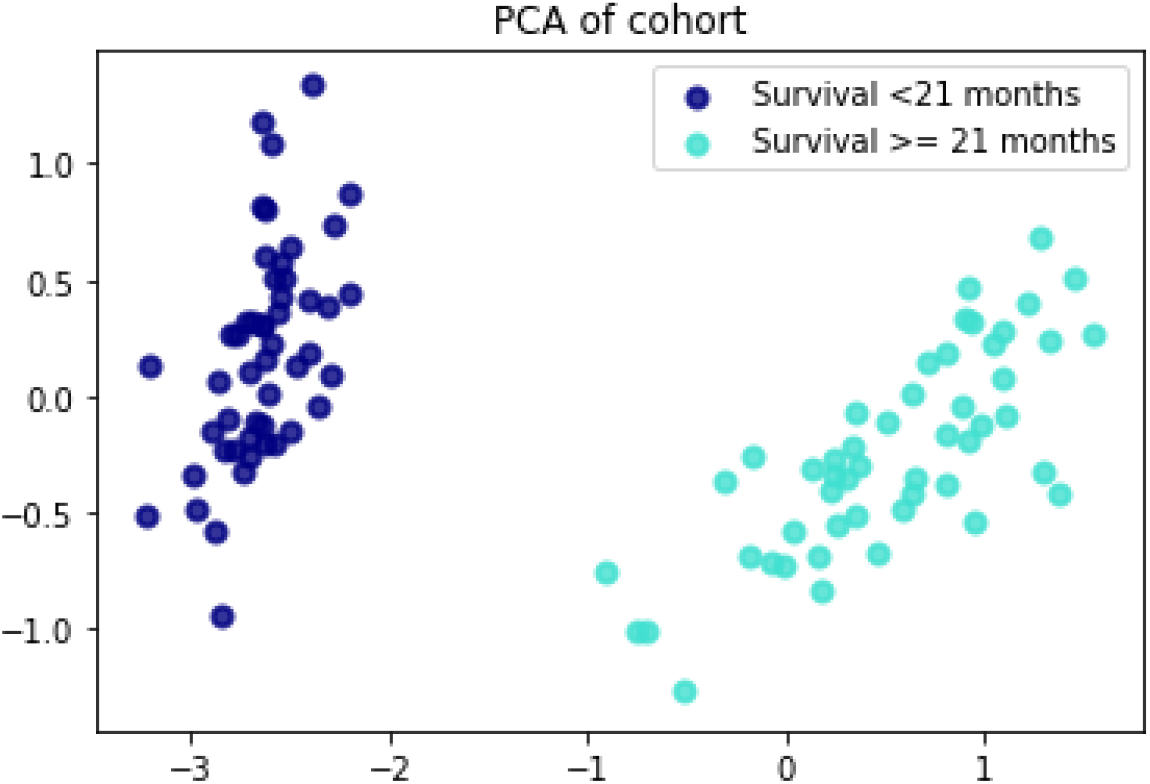
PCA plot of patients. Patients surviving less than XX years are shown in red; otherwise green.

### Predictive Model Performance Metrics

Across all folds of cross validation, the model achieved AUC of 0.649, accuracy of 0.752, precision of 0.623, recall of 0.051, and F1 score 0.094. The receiver operating curve is shown in figure 3.

**FIgure 3:**
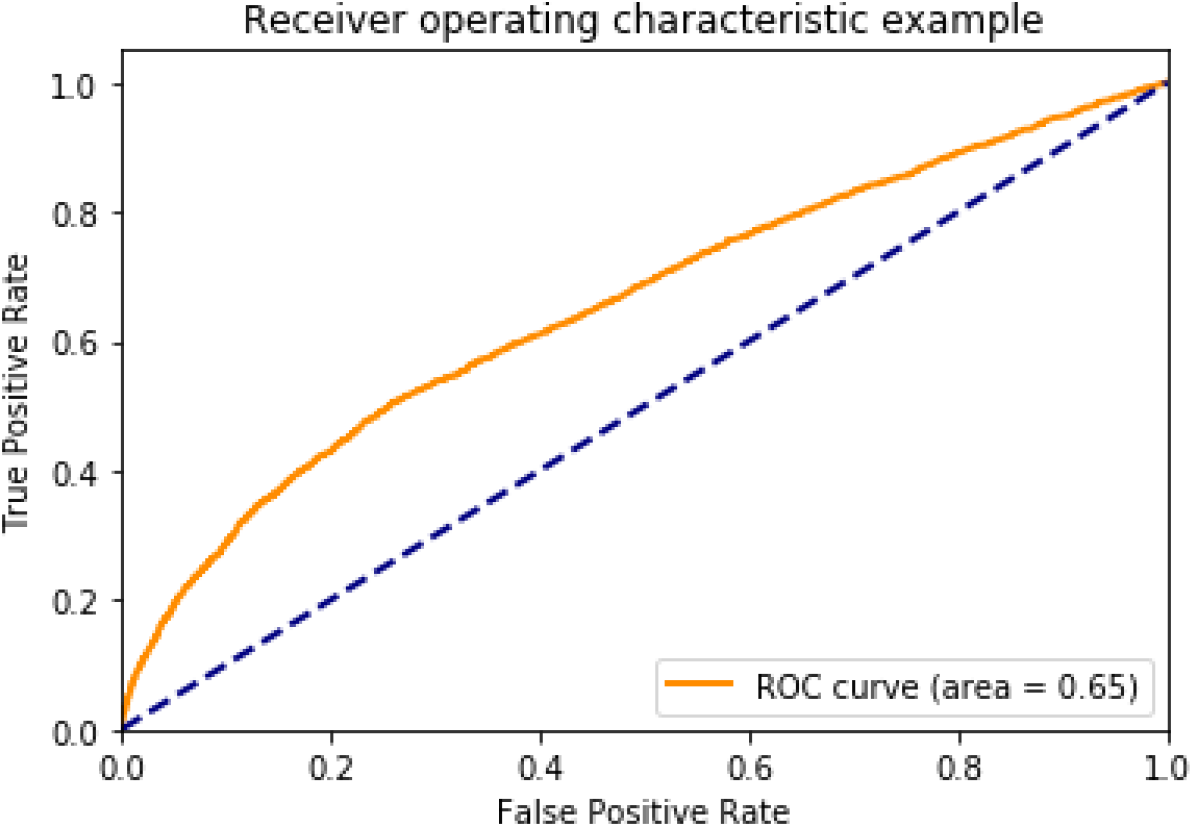
Receiver operating curve for the logistic regression model.

### Feature Importance

Of the 19 most predictive features with non-zero weights learned from the logistic regression model, all of them conveyed clinical meaningfulness in the application of early mortality prediction. Metastases to the bone, lung, liver and brain were positively predictive of early mortality, with bone being the metastatic site with strongest clinical predictiveness. Features are visualized in terms of predicted weights in Figure 4.

**Figure 4:**
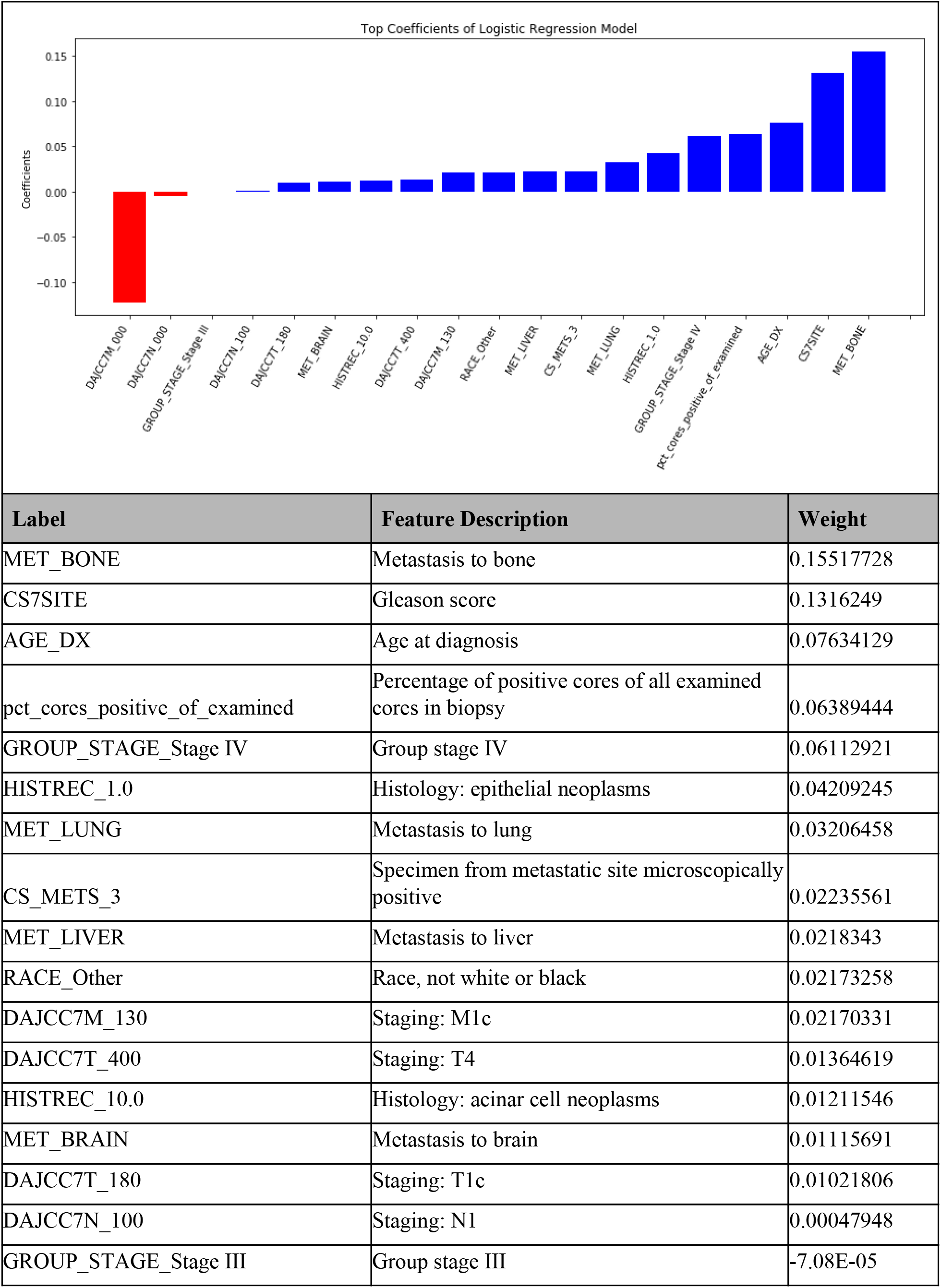
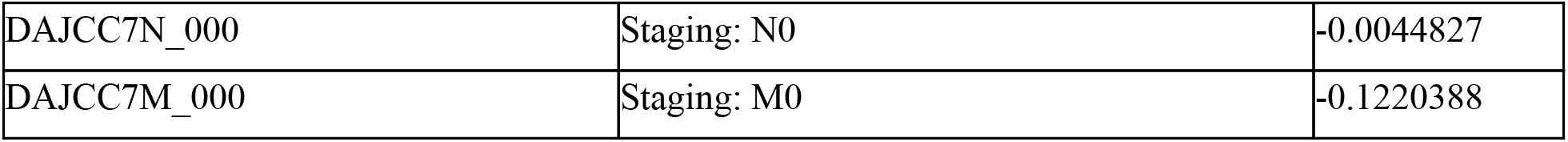
top most predictive features, including all features with a positive weight or negative weight learned from the model. Positive weights indicate positive correlation with early mortality, while negative weights indicate negative correlation with early mortality.

## Discussion

In this study, we built a machine learning model to predict early mortality (less than 21 months, the median overall survival of patients with stage III and IV prostate cancer).

The most predictive features were arrived at via extraction of weights learned from the logistic regression model. It is important to note that the most predictive features were determined from feature coefficients in the model. In our logistic regression model, feature importances were interpreted based on coefficients learned from the model. In clinical applications, interpretability is important for downstream usage of the model in personalized treatment plans for patients. Methods such as LIME [17] have been successfully used in healthcare applications including prediction of mortality in ICU patients [18].

It is important to note that there are methods for identifying risk factors as correlated to mortality, such as Cox proportional hazards model. We did not implement a Cox model in this scenario because the problem was setup as a prediction problem.

There are three main areas for future work. First, the incorporation of additional features such as medications, comorbidities, behavioral factors (e.g., ECOG), genomics, insurance type, health system encounters (e.g., inpatient encounters, hospitalizations) should be explored. Prior work on machine learning for early detection of disease has shown that inpatient encounters have been strongly predictive of early mortality. [5] While the SEER database does not include such features, a study could be performed from electronic health records (EHRs) as a data source.

The second area of future work involves leveraging temporal models that take into account the relative temporal relationship between occurrences of features. Models that take into account temporal relationships between features include autoencoders for learning phenotypic features from temporal features such as labs[19], as well as recurrent neural network models which learn predictive representations that capture temporal relationships between features[20].

The third area of future work involves validation of the model trained on SEER data, in other data sources. Transfer learning methods can be employed. There are various ongoing efforts of standardization of EHR data models such as Health Level 7 (HL7) standard Fast Healthcare Interoperability Resources (FHIR) [21]and Observational Health Data Sciences and Informatics (OHDSI): opportunities for observational researchers (OHDSI)[22], which will enable improved reproducibility of scientific research.

## Conclusion

We developed a machine learning model that predicts early mortality in prostate cancer patients with 75.2% accuracy with AUC 0.649, and is able to identify clinical features predictive of early mortality.

Future work should include integration of additional data sources, as well as explore temporal modeling strategies to account for clinical changes over time.

## Data Availability

Public data was used.

https://seer.cancer.gov/

## Footnotes

1 See NCI SEER definition: https://training.seer.cancer.gov/schema/rp_ureter/reg_ln_eval.html

2 See NCI SEER definition: https://staging.seer.cancer.gov/cs/input/02.05.50/prostate/mets_eval/?breadcrumbs=(~schema_list~),(~view_schema~,~prostate~)

